# Intervention and evaluation protocol of *fit4future Kids*: A multi-component health promotion programme in German primary schools

**DOI:** 10.64898/2026.05.23.26353928

**Authors:** Simon Blaschke, Katharina Sterr, Daniela Heß, Lina Lux, Anna Brandmeier, Filip Mess

**Affiliations:** Professorship of Didactics in Sport and Health, School of Medicine and Health, Technical University Munich; Institute for Movement Therapy and Movement-oriented Prevention and Rehabilitation, German Sport University Cologne

## Abstract

**Background:** Schools are widely recognised as key settings for promoting children’s health behaviours. However, many schools struggle with the implementation and especially sustainment of health promotion programmes e.g. due to limited resources. Strengthening schools’ capacity for health promotion has therefore been identified as a central strategy for achieving better implementation and ultimately behaviour change outcomes among children. The fit4future Kids programme was developed as a large-scale, multi-component initiative in Germany that aims to promote children’s physical activity, nutrition, mental health, and responsible digital media use while simultaneously supporting schools in building structures for sustainable health promotion.

**Methods:** This paper describes the intervention and evaluation protocol of the nationwide fit4future Kids programme implemented in several cohorts of German primary schools from Sept. 2022 to Sept. 2027. The intervention is based on the Health Promoting Schools framework and integrates established implementation and behaviour change frameworks, including the Consolidated Framework for Implementation Research, the COM-B model, and Behaviour Change Techniques. The programme combines curricular materials, environmental components, and structured implementation support to facilitate the integration of health promotion into everyday school practice. The evaluation follows a mixed-methods design involving multiple stakeholder groups, including school staff, parents, and children. Quantitative and qualitative data are collected to assess implementation processes, contextual factors, and programme outcomes. The large and diverse sample of 1,153 participating primary schools allows for the exploration of different implementation trajectories and the investigation of potential equity-related effects.

**Discussion:** By combining evidence-based health promotion strategies with implementation science approaches, fit4future Kids provides a large-scale real-world example of how schools can be supported in implementing sustainable health promotion. The evaluation is expected to generate important insights into the implementation and potential effectiveness of multi-component school-based interventions and to inform future initiatives aiming to strengthen health-promoting school environments.

## Introduction

### Relevance

The prevalence of non-communicable diseases (NCDs) has risen substantially over the past decades, becoming the leading cause of death and disability-adjusted life years worldwide [1]. Annually, NCDs account for over 40 million deaths, representing 71% of all deaths globally [2]. Health behaviours play a critical role in NCD risk, with approximately 29% of deaths linked to dietary factors such as low consumption of whole grains and fruits [3], and around 8% attributed to insufficient physical activity [4]. Additionally, behaviours related to media use and stress management contribute indirectly by interacting with other risk factors and exacerbating NCD progression. Consequently, addressing multiple lifestyle factors including diet, physical activity, stress management, and media use simultaneously appears essential for effective NCD prevention [5].

While these lifestyle factors are important targets, evidence highlights the critical importance of intervening during childhood to effectively reduce NCD risk [6–9]. Studies demonstrate that risky health behaviours and elevated stress levels in childhood are associated with increased NCD risk later in life [10,11]. The developmental window of middle childhood (ages 6 to 12 years) is particularly pivotal, as lifestyle habits consolidated during this period often persist into adolescence and adulthood [12]. However, data from a German population representative study on school children reveal concerning prevalence rates: only about 11% of girls and 21% of boys meet recommended physical activity guidelines [13]. Regarding dietary behaviour in primary school children, merely 7% meet WHO recommendations of limiting free sugar intake to less than 10% of daily energy, while 80% of girls and 83% of boys exceed this threshold by at least 1.5 times [14]. Furthermore, only 14% of children in this age group consume the recommended portions of fruit [15]. The longitudinal COPSY study reports that approximately 28% of children in Germany experience mental health problems, with 45% of adolescents facing mental stress and 49% reporting exhaustion or fatigue at least weekly [16,17]. Socioeconomic status (SES) further influences these health behaviours and NCD prevalence [18,19]. Among German children, lower SES correlates with unhealthy dietary patterns, reduced physical activity, increased media use, and higher risk of mental health issues [20,21]. Low SES also impedes the effectiveness of health promotion interventions due to barriers such as limited access to health services, time constraints, and financial hardship [19].

To reduce these inequities, the school setting provides a promising platform to reach children regardless of SES and deliver sustainable health promotion [19,22–24]. Primary schools in Germany offer access to large numbers of children in their daily environments, enabling systematic and sustainable health promotion integrated into routine activities [25]. The WHO’s Health Promoting School (HPS) framework embodies this approach by emphasizing individual behaviour change alongside supportive school environments and strong community linkages. An HPS is defined as a school continually enhancing its capacity to be a healthy place for living, learning, and working [26]. Accordingly, multicomponent health promotion programmes targeting multiple lifestyle behaviours simultaneously at individual and environmental levels have strong potential to reduce NCD risk during this critical developmental stage and across socioeconomic groups [27].

### Theoretical background

Evidence supports the positive effects of school-based health promotion using the setting approach on children’s health behaviours [19,23,28]. Specifically, interventions in primary schools addressing physical activity, nutrition, media use, and mental health yield small to moderate beneficial effects on NCD risk factors [29,30]. Reviews recommend multicomponent approaches combining physical activity, diet, and stress coping strategies to maximise effectiveness [31,32]. Factors that can in addition enhance effectiveness include capacity building within schools, such as engaging all relevant school stakeholders [33], direct support from health promotion experts [34], and fostering school networks [35], and teacher training to improve implementation [36] as well as programme design, e.g. tailored programme components [37] and blended digital and in-person delivery [38]. However, concerning the effectiveness of these programmes, most studies focus on short- and mid-term outcomes, with a scarcity of long-term evaluations beyond six months [27]. The sustainability of school-based health promotion is also questioned in a review by Herlitz et al., which found that none of 18 programmes were fully implemented in the long term, with only some components maintained post-intervention [39]. Social inequities compound these challenges, as resource limitations and infrastructure deficits in disadvantaged areas hinder implementation success [19]. These findings underscore the need to systematically address implementation factors to fully exploit the potential of school health promotion.

This is where implementation science might come into play [40]. According to Moir [41], implementation science “is concerned with using a systematic and scientific approach to identify the range of factors which are likely to facilitate the administration of an intervention” (p.2). Further, “Implementation science focuses on developing and testing methods to broadly spread successful sustained implementations across diverse settings” [42] (p.1). Originally emerging from the clinical field, implementation science has now extended into various settings, where health promotion and prevention takes place. This expansion has been facilitated by the development of frameworks, which not only broaden the applicability of implementation science beyond clinical contexts but also render it empirically testable [42]. In its latest version, the Consolidated Framework for Implementation Research (CFIR) offers a comprehensive framework for implementation planning and evaluation [43–45]. It encompasses determinants, strategies and outcomes and provides terminology, which we will use as much as possible throughout this protocol paper. While the CFIR only covers some implementation strategies, the Expert Recommendations for Implementing Change (ERIC) provides an extensive set of evidence-based implementation strategies tailored to healthcare contexts [46]. To translate these strategies for school health promotion, Cook et al. developed the School Implementation Strategies Translating ERIC Resources (SISTER) framework, identifying 75 strategies grouped into nine categories, rated as relevant for the school context by experts in the field [47]. Examples include strategies such as providing interactive teacher training in the category of training and educating stakeholders [47]. Damschroder et al. recommend extending the CFIR with these taxonomies by linking determinants and strategies [44]. While these evidence-based frameworks can improve implementation planning and evaluation, data on their application in real-world settings remain limited [48,49]. Addressing this limitation in current research and further specifying the mechanisms through which programme activities are expected to influence behaviour is facilitated by the COM-B model [50]. This model is integrated into the CFIR by making explicit how intervention content and implementation strategies are linked to behavioural determinants and is further enhanced using Behaviour Change Techniques (BCTs), which provide a systematic taxonomy for specifying the active ingredients of the intervention [51].

### The fit4future Kids programme

Based on these advances in current literature, the *fit4future Kids* health promotion programme exemplifies a primary school-based, multicomponent, capacity-building and partially tailored blended intervention incorporating a comprehensive concept for implementation and evaluation relying on the CFIR and recommended extensions (SISTER, COM-B framework, BCTs). Concerning the programme content, the intervention targets physical activity, nutrition, mental health and digital media use, based on the requirements set out by the German Prevention Guideline [52].

Previous versions of this programme were partially evaluated and demonstrated effectiveness on physical activity and functioning [53], but similar to other health promotion programmes in primary schools lacked systematic implementation guidance and follow-up evaluation [27,39]. Therefore, the updated programme addresses these gaps by mapping each programme component onto the COM-B model to clarify whether it primarily targeted capability, opportunity, or motivation among the involved stakeholders. To further increase transparency and comparability with international research, the updated programme selects components in line with the BCTs [51], thereby specifying the active ingredients through which change is intended to occur. Recent studies further highlight the added value of linking implementation strategies with BCTs and COM-B, as this combination helps to elucidate mechanisms of change and clarify how strategies influence determinants and outcomes [54–56]. As another novelty, the updated programme builds schools’ capacity for implementation by integrating the Plan–Do–Study–Act (PDSA) cycle within the SISTER taxonomy of implementation strategies. This strategy is supported by studies, such as Tichnor-Wagner et al., stating that educators in public school districts valued PDSA cycles for continuous improvement and that PDSA training helped build implementation capacity [57].

This protocol details the *fit4future Kids* programme, integrating current evidence on effective school health promotion and implementation science. It outlines the programme’s aims and overview, the programmes partners, and gives a detailed description of its procedure, intervention (innovation) and implementation components, as well as evaluation plan. Thus, the Template for Intervention Description and Replication (TIDieR) checklist to enhance transparency and replicability will be used to describe *fit4future Kids* (see S0) [58]. Beyond programme and evaluation description, this protocol constitutes a transferable methodological resource that supports best practices and evidence-informed design of sustainable school-based health promotion initiatives at an international level.

## Methods

### Programme aim

With the theoretical background outlined in the introduction, the *fit4future Kids* programme aims to achieve two main goals: (1) the advancement of health and health-promoting behaviours in primary school children, and (2) building schools’ capacity to sustainably and continuously implement health-promoting activities in line with the HPS framework.

### Overview

*fit4future Kids* is being implemented over four years, from September 2022 to December 2026. It targets children aged 6 to 12 years attending primary schools in Germany, as well as school personnel and parents or caregivers as key stakeholders. While primary education in Germany typically covers children up to approximately 10 years of age, the upper age range reflects variation across federal states, where primary schooling may extend to six years in some regions. The programme is designed to reach a large and diverse population and is implemented nationwide, with particular emphasis on including primary schools from socioeconomically disadvantaged regions to enable equity in health promotion.

The programme is structured into successive cohorts, with the first cohort starting in the 2022/2023 school year. Each following school year, additional schools are enrolled and assigned to their respective cohort. Cohort 1 participates for four years, cohort 2 for three years beginning in 2023/2024, and cohort 3 for three years starting in 2024/2025. Recruitment for cohorts 1-3 has been completed, but data collection is still going on for all three cohorts. An overview is provided in Fig. 1. Throughout the programme’s implementation, continuous feedback and lessons learned are systematically collected and integrated to optimize the programme’s implementation and effectiveness, which will be described in detail in the evaluation section. Based on this iterative process, the programme was previously refined and condensed into a version of the programme with a duration of two years, which constitutes the intervention in cohort 3 in the 2024/2025 school years. Fig 2 shows the overall structure of the programme.

**Fig 1.**
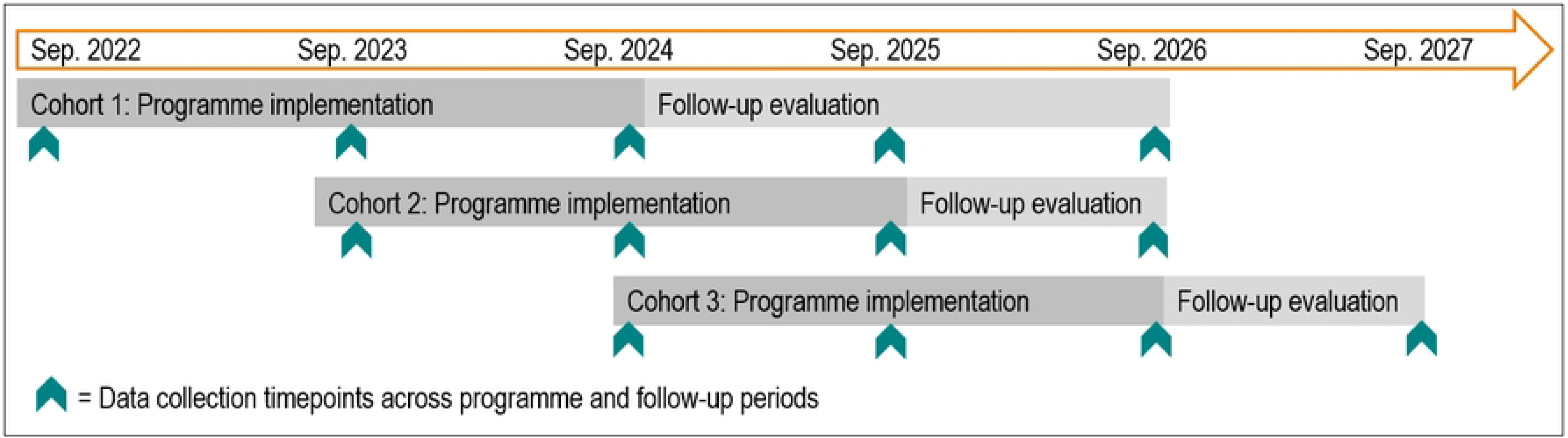
Cohort overview.

**Fig 2.**
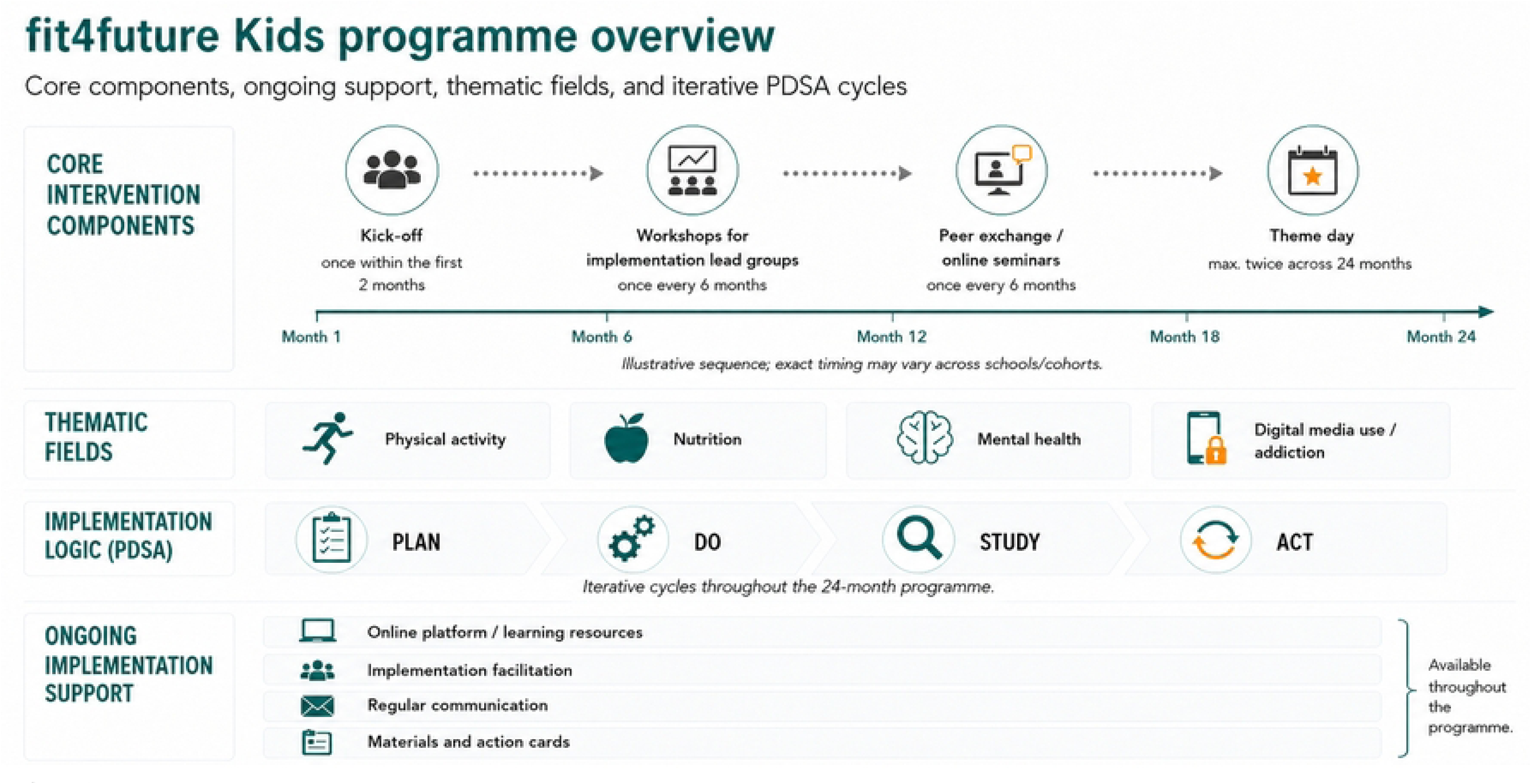
Programme structure overview.

### Programme partners

The overall programme development, organisation, and management is carried out by *planero GmbH*, which is a company that develops and provides health promotion programmes primarily in the school setting. The intervention content was developed collaboratively by the Professorship of Preventive Sports Medicine and Sports Cardiology at the Technical University of Munich (TUM) in cooperation with *fischimwasser GmbH*. *fischimwasser GmbH* develops evidence-based solutions to promote healthy lifestyles and sustainable well-being in different settings. The evaluation of *fit4future Kids* is conducted by the Professorship of Sport and Health Didactics at TUM. The programme is funded by the health insurance provider *DAK Gesundheit*.

### Programme description

The implementation process of *fit4future Kids* follows a structured sequence from school recruitment to provision of materials, training, and sustained support. While some components are mandatory, schools have a high degree of flexibility in how they tailor the programme to their specific needs. The narrative below outlines the chronological flow and content of components, with each systematically described in terms of its underlying COM-B mechanisms, the BCTs through which these mechanisms are targeted, and the corresponding implementation strategies (classified via SISTER). We present components in this order: COM-B, BCTs, and SISTER strategies to move from mechanisms, to active ingredients, to implementation strategies, ensuring conceptual clarity and alignment with recent advances in implementation research [54–56]. A comprehensive overview of the mapping is provided in S1.

#### Recruitment

To ensure representativeness and equity, recruitment was designed to achieve a balanced inclusion of schools with and without prior health promotion experience, and approximately 25% of schools from socially disadvantaged contexts [59]. To identify socially disadvantaged schools, SINUS-Milieus data [60] was used as a proxy for local socioeconomic conditions. Specifically, the concentration of the precarious milieu within a 1 km radius of each school was calculated and normalized against regional averages to account for structural demographic differences [61].

Recruitment took place via official education authorities, existing regional health and education networks, and proactive outreach by the programme team. Education authorities were initially contacted via email and, where appropriate, invited to informational video meetings to introduce the programme in detail. Schools and other educational institutions were approached through multiple channels, including email contact by *planero GmbH*, follow-up telephone calls, and, in some cases, written invitations. In addition to network-based recruitment, cold acquisition via telephone was employed to reach schools beyond established contacts and to ensure regional diversity. Recruitment efforts were further supported by website-based promotion through the *DAK-Gesundheit* and by the distribution of informational flyers at relevant conferences and fairs.

Participation was confirmed by school principals through a digital registration form, after which schools received structured onboarding support and successive access to programme materials.

Across three cohorts, 1,153 primary schools were recruited, which represents around 9% of all primary schools in Germany [62]. This exceeds common sampling percentages used at the acquisition stage for designing representative studies [63]. A comparable programme reported an 8% response among contacted schools [64], indicating that the targeted scale was feasible for large school-based health promotion. 619 schools participated in cohort 1, 251 in cohort 2, and 283 in cohort 3 (Table 1).

**Table 1.**
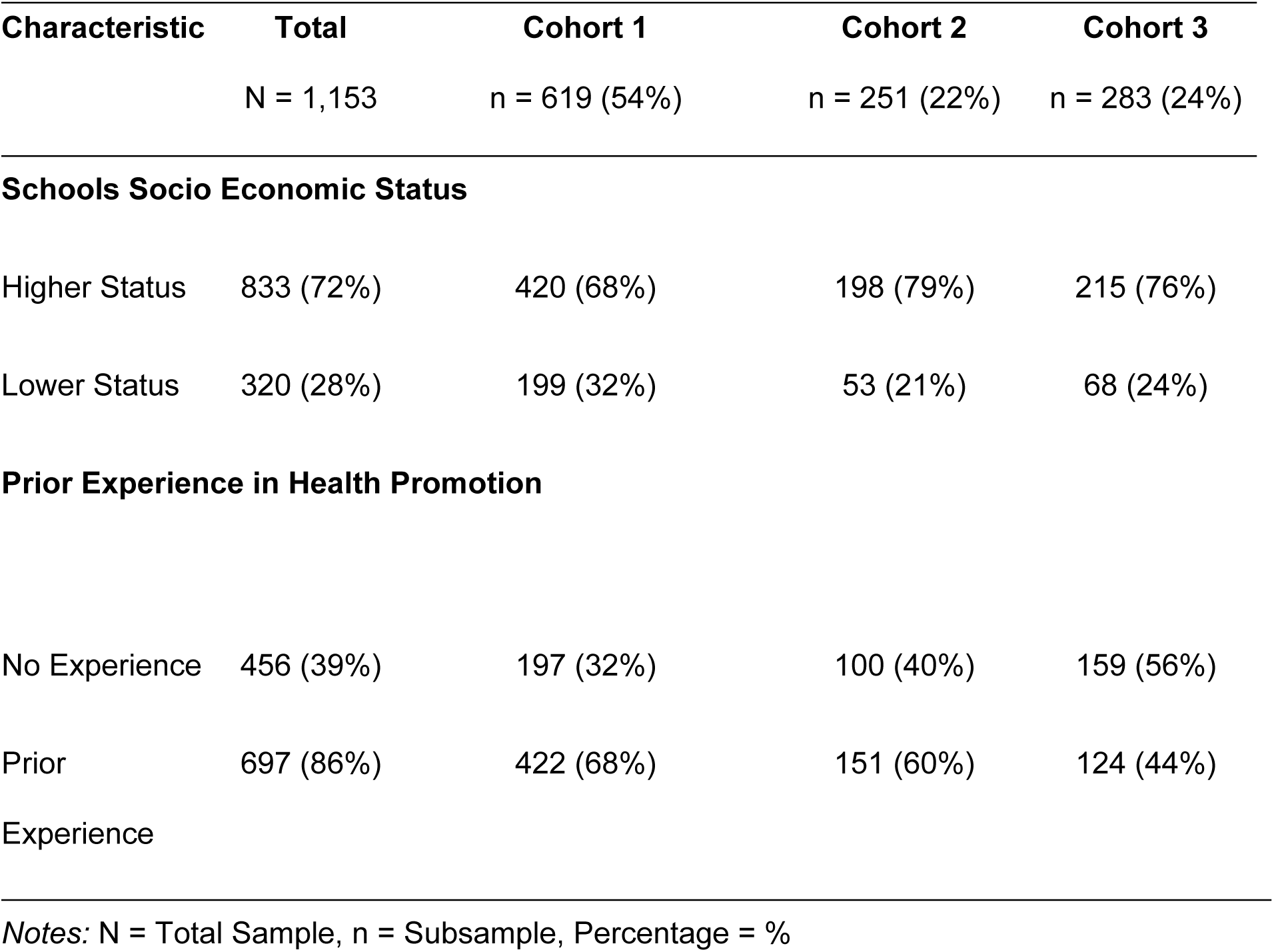
Sample Characteristics of Primary Schools by Cohort.

| Characteristic | Total | Cohort 1 | Cohort 2 | Cohort 3 |
| --- | --- | --- | --- | --- |
|  | N = 1,153 | n = 619 (54%) | n = 251 (22%) | n = 283 (24%) |
| **Schools Socio Economic Status** |  |  |  |  |
| Higher Status | 833 (72%) | 420 (68%) | 198 (79%) | 215 (76%) |
| Lower Status | 320 (28%) | 199 (32%) | 53 (21%) | 68 (24%) |
| **Prior Experience in Health Promotion** |  |  |  |  |
| No Experience | 456 (39%) | 197 (32%) | 100 (40%) | 159 (56%) |
| Prior Experience | 697 (86%) | 422 (68%) | 151 (60%) | 124 (44%) |

Notes: N = Total Sample, n = Subsample, Percentage = %

#### Assignment of implementation facilitators

After recruitment, each participating school was assigned an external implementation facilitator (called Area Manager in the programme) who provided ongoing support to several schools within one geographical region. Implementation facilitators were trained by programme developers, namely *planero GmbH* and *fischimwasser GmbH*, combining role-specific competence and content-related training. Training was delivered in four one-day sessions over two years with semi-annual follow-up until programme completion. Its overall aim was to clarify facilitators’ role understanding and ensure good implementation practice [65]. We deliberately included the coaching literature, as it offers more established role definitions and structured competence frameworks compared to the still emerging concept of implementation facilitators [66].

For role-specific competence training, a literature review identified frameworks from implementation facilitation, coaching, and health coaching [65,67–70]. The resulting competence profile aligned with the coach education model [71], covers personal, social-communicative, professional-methodological, and action competence and guided the development of a process-oriented training programme integrating knowledge transfer, practice, and reflection. The four modules of this training programme addressed (1) competencies, motivation types, and communication, (2) structuring and self-management, (3) motivational communication and resistance using motivational interviewing techniques, and (4) autonomy and independence, including practice-oriented methods such as confidence talks [72]. This modular design supported continuous role-specific training, professionalization, and preparation for the diverse demands of programme implementation in the school setting. An overview of the implementation facilitator training is given in S2.

With regard to content-specific training, implementation facilitators were qualified health professionals with formative education backgrounds in nutrition, physical activity, mental health, and media use. Their role was to convey programme content to the implementation lead groups (see next chapter) in workshops. For this purpose, they received workshop materials in advance, worked through them in groups, and presented sections to their peers with subsequent feedback and input. This approach fostered exchange of expertise, strengthened knowledge transfer skills, and prepared facilitators to address potential critical questions. Training was supervised by the programme developers to ensure fidelity and consistent competence across implementation facilitators and schools. Within the COM-B framework, implementation facilitators primarily enhance psychological capability by building knowledge, skills, and role clarity through training as well as social opportunity through their continuous support and function as external change agents [73]. They also contribute to reflective motivation by modelling good practice and maintaining engagement over time. Relevant BCTs include *instruction on how to perform the behaviour*, *demonstration of the behaviour*, *feedback and monitoring*, *social support (practical)*, and *problem solving*. From a SISTER perspective, this corresponds to strategies such as *Train and educate stakeholders* (through structured training modules), *Provide interactive assistance* (ongoing support to schools), and *Develop stakeholder interrelationships* (acting as a bridge between schools and the programme team).

#### Establishment of an implementation lead group

Each participating school formed its own implementation lead group, consisting of members of the school leadership (e.g. the principal), two teachers or school social workers, and one parent representative. This group was responsible for coordinating programme communication, participating in live trainings, and embedding activities within school structures [74]. Within the COM-B framework, this restructuring of the social environment primarily enhanced social opportunity and reflective motivation, supported by BCTs such as *social support, action planning,* and *involvement of credible sources*. From an implementation strategy perspective, this aligns with the SISTER category *Develop stakeholder interrelationships*. Through shared responsibility within the implementation lead group and the resulting school-level social structure, reflective motivation and social opportunity for implementing change are expected to increase.

#### Access to the digital platform

Implementation lead groups received access to the *fit4future Kids* platform, the central online hub for materials and communication, comprising: (1) Idea Box (digital repository; S3), (2) Blog (How-To, Best-Practice, inspirational posts; S4), (3) Useful Information (programme-level materials and PDSA tools by cohort; communication and evaluation resources; S5), and (4) Events (planned/past events). Within the COM-B framework, the platform primarily strengthens psychological capability (knowledge, skills, health literacy), complemented by reflective motivation (inspirational content) and social opportunity (exchange formats). The related BCTs include *instruction, information provision, modelling, prompts*, and *social support*. From a SISTER perspective, the platform represents strategies such as *Train and educate stakeholders* and *Develop and distribute educational materials*, and, through its tools, also contributes to *Use evaluative and iterative strategies*.

#### Kick-off and provision of materials

Following registration and the assignment of an implementation facilitator, schools participated in an online kick-off meeting that introduced the goals, structure, and processes of the programme. This initial orientation aimed to create a shared understanding across schools and establish motivation and commitment among school teams.

Within the first weeks of implementation, each school received the *fit4future Kids* box (S6), containing play and sports equipment as well as action cards (S7) designed to catalyze active routines in the school day. The materials provided ready-to-use opportunities for integrating short activity breaks, games, and movement-based learning into everyday practice.

From a COM-B perspective, the kick-off meeting primarily addressed reflective motivation (through goal-setting, shared vision, and orientation), while the box and action cards strengthened physical and psychological capability (by equipping schools with the knowledge and tangible resources needed to initiate change) and physical opportunity (providing equipment to enable activity). Relevant BCTs include *goal setting (behaviour)*, *instruction on how to perform the behaviour*, *adding objects to the environment*, and *prompts/cues*. From an implementation strategy perspective (SISTER), these components correspond to *Train and educate stakeholders* (kick-off), *Provide interactive assistance*, and *Develop and distribute educational materials* (*fit4future Kids* box).

#### Workshops for implementation lead group

In-person or digital workshops targeted members of each school’s implementation lead group. Co-developed with scientific partners and delivered by implementation facilitators in structured learning groups, each workshop focused on one core topic (physical activity, nutrition, mental health, or digital media use/addiction) aligned with the German Prevention Guideline [52]. To simultaneously strengthen schools’ implementation capacity, each workshop also emphasized one step of the PDSA cycle. For example, the first workshop combined content on physical activity and active breaks during the school day with an introduction to the planning phase of the PDSA cycle. Designed as the programme’s central intervention component, the workshops aimed to build both health-related competencies and process knowledge for independent implementation. Within the COM-B framework, these workshops primarily enhanced psychological capability, e.g. knowledge, skills and health literacy of the implementation lead group and were complemented by reflective motivation through structured goal-setting and practical relevance. Relevant BCTs include *instruction on how to perform the behaviour*, *behavioural practice/rehearsal*, *action planning*, and *problem solving*. From a SISTER perspective, the workshops represent strategies such as *Train and educate stakeholders*, *Develop stakeholder interrelationships*, and *Use evaluative and iterative strategies* through the application of PDSA cycles. Exemplary material from the workshops is displayed in S8.

#### Workshops for all stakeholders

These digital events (called *Online Seminars* in the programme) are held twice a year by external topic experts and are open to all teachers and parents or caregivers, regardless of cohort, lead group membership, or prior experience. Implementation facilitators provided additional, workshop-related support where needed, for example by helping schools adapt materials to their local context, clarifying roles and next steps for the implementation lead group, and assisting with the planning of initial activities at the school [33]. The seminars addressed the programme’s core topics of physical activity, nutrition, mental health, and digital media with the aim of deepening knowledge and competencies beyond the content of the lead group workshops [52]. Within the COM-B framework, these seminars primarily enhance psychological capability (knowledge and skills) and reflective motivation (through exposure to expert input and broader relevance). Relevant BCTs include *information about health consequences*, *instruction on how to perform the behaviour*, and *credible source*. From a SISTER perspective, the seminars reflect *Train and educate stakeholders* and *Engage consumers* by directly involving parents or caregivers alongside school staff.

#### Peer-to-peer exchange sessions

These digital meetings (in the programme called *Online Get-Togethers*) are led by the implementation facilitators. They provide space for schools to exchange experiences and build networks. Peer-to-peer exchange sessions are directly connected to the workshop content and tasks, additionally supporting schools in applying the PDSA cycle. Within the COM-B framework, peer-to-peer exchange primarily enhances social opportunity through structured interaction and reflective motivation by normalizing change efforts and providing mutual encouragement. Relevant BCTs include *social support (practical/emotional)*, *restructuring the social environment*, and *problem solving*. From a SISTER perspective, these sessions represent *Develop stakeholder interrelationships* and *Provide interactive assistance* through facilitated peer learning.

#### On-site Health Activity Day

Schools were given the opportunity to organise a Health Activity Day in collaboration with their implementation facilitator. These on-site events involve children, parents or caregivers, and teachers and offer experiential learning opportunities across the four core topics (S9). If needed, schools may also request individual coaching by the implementation facilitator to receive tailored support in addressing specific challenges. Within the COM-B framework, Health Activity Days primarily enhance physical opportunity (by creating structured occasions for activity) and psychological capability (through hands-on learning and experiential practice). They may also contribute to reflective motivation by engaging the wider school community and highlighting relevance across contexts. Relevant BCTs include *adding objects to the environment*, *demonstration of the behaviour*, *behavioural practice/rehearsal*, and *social support (practical/emotional)*. From a SISTER perspective, Health Activity Days represent strategies such as *Develop and distribute educational materials* (through resources provided), *Engage consumers* (by involving parents or caregivers and students), and *Tailor strategies to context* (via individualized coaching where needed).

#### Ongoing support

Throughout the programme, schools receive continuous support from their assigned implementation facilitators and the programme team. Regular reminder and impulse mails help sustain engagement. Within the COM-B framework, ongoing support primarily strengthens reflective motivation (through prompts and encouragement) and social opportunity (through continuous access to facilitator guidance). Relevant BCTs include *prompts/cues*, *feedback and monitoring*, and *social support (practical/emotional)*. From a SISTER perspective, ongoing support corresponds to *Provide interactive assistance* and *Use evaluative and iterative strategies* by maintaining contact and adapting impulses to schools’ needs.

#### Incentives and certification

To acknowledge the engagement of implementation lead group members, school staff who complete the required number of workshops and peer-to-peer exchange sessions during the programme receive a process facilitator certificate. This certificate accredits teachers as health promotion experts within their schools and counts toward mandatory professional development. Importantly, all programme participation, materials, and events are free of charge for schools, as the programme is fully funded by a statutory health insurance provider, which itself serves as an additional incentive by lowering barriers to participation. Within the COM-B framework, certification and free access primarily enhance reflective motivation (through recognition and reduced cost barriers) and social opportunity (by strengthening the formal role of health promotion experts within schools). Relevant BCTs include *material incentive (behaviour)*, *social reward*, *credible source*, and *remove financial barriers*. From a SISTER perspective, these components correspond to *Use incentives* and *Engage consumers/stakeholders* by linking programme participation with formal recognition and free access to high-quality resources.

#### Tailoring of programme components

Overall, the programme was designed to allow flexible application and adaptation according to the needs and priorities of individual schools. While several core components were mandatory to ensure a common implementation structure across schools, including the establishment of an implementation lead group and participation in implementation lead group workshops, other components were optional such as peer-to-peer exchange sessions, Health Activity Days, and additional coaching sessions with implementation facilitators. This flexible structure aimed to provide schools with a high degree of autonomy while maintaining a shared implementation framework. Adaptations and decisions regarding the use of programme components were encouraged to be reflected within the PDSA cycle to support context-sensitive but structured implementation and continuous improvement processes. Within the COM-B framework, this tailoring approach primarily supports motivation by increasing autonomy, ownership, and perceived relevance of programme activities, while also strengthening opportunity by improving the fit between programme components and local school priorities. From a BCT perspective, this approach aligns most closely with *problem solving*, as schools were encouraged to continuously reflect on their individual implementation challenges and identify context-sensitive solutions through the PDSA cycle. From a SISTER perspective, this corresponds particularly to *Tailor strategies*.

### Programme evaluation

#### Evaluation aims and design

Aligned with the two programme aims of *fit4future Kids*, the evaluation examines both implementation and effectiveness using a mixed-methods approach at the levels of the implementation lead group, parents or caregivers and implementation facilitators.

At the implementation lead group level, the evaluation focuses on the programme’s aim of building schools’ capacity to sustainably and continuously implement health-promoting activities in line with the HPS framework. More specifically, the aim is to evaluate how *fit4future Kids* is embedded into school structures through group interviews to assess overall acceptability and feasibility, investigate barriers and facilitators to the adoption and implementation of health-promoting activities, as well as the sustainment of programme components. These group interviews are conducted annually in cohort 2 (starting in 2023) and cohort 3 (starting in 2024) with the implementation lead group, resulting in three group interview time points. Cohort 1 was not included in this component of the evaluation due to temporal overlap between the baseline assessment and programme initiation, as well as financial constraints. Furthermore, the evaluation on this level examines for example, perceived environmental and personal health promotion practices at the respective school as well as the schools’ status in applying the PDSA cycle for health-promoting initiatives and schools’ organisational health literacy by collecting survey-based data. This survey is employed with cohort 1 (starting in 2022), at the beginning of every school year which results in five measurement time points for cohort 1, four measurement time points in cohort 2 (starting in 2023) and four measurement time points in cohort 3 (starting in 2024). Additionally, the implementation lead group fills out online feedback questionnaires after the onboarding workshops as well as all other workshops, seminars and peer-to-peer exchange sessions to examine appropriateness, acceptability and feasibility of the respective programme content. Lastly, the implementation lead groups’ usage of the programme is measured by tracking the amount of digital platform logins and participation in workshops and peer-to-peer exchange sessions as objective outcomes of programme implementation.

At the parental or caregiver level, the evaluation addresses the programme aim of advancing health and health-promoting behaviours in primary school children. Thereby it also aims to assess implementation by quantifying, among other aspects, usage and acceptability of the programme and its components and examine effectiveness with respect to, for example children’s health and health behaviours as well as parental health literacy and the family health climate within an online survey. This online survey is employed only in cohort 2 (starting in 2023) with parents or caregivers assessing the constructs at the beginning of the programme enrollment, which was aligned with the start of the school year, after one year and after two years resulting in three measurement time points for these evaluation components.

At the implementation facilitator level, the evaluation concerns an external perspective on programme delivery and school development. Annual semi-structured focus group interviews with implementation facilitators (starting in summers 2024) explore (1) usage of the *fit4future Kids* components and the underlying reasons, (2) application of the PDSA cycle, (3) observable changes in schools’ health-promoting activities and organisational processes, and (4) demands for additional support communicated by the implementation lead group in the programme implementation and how this was provided by implementation facilitators. Furthermore, implementation facilitators also fill out feedback surveys after receiving programme training or delivering programme content as programme experts.

To address these aims, the evaluation design was developed by the Professorship of Sport and Health Didactics at TUM in cooperation with *planero GmbH* and *fischimwasser GmbH*, which were involved in evaluation organisation or minor evaluation aspects. In line with current guidance for implementation and effectiveness evaluations, the programme evaluation draws in particular on the updated CFIR [44], Proctor’s Implementation Outcomes Framework [75] and the Reach, Effectiveness, Adoption, Implementation, and Maintenance framework (RE-AIM) [76,77]. Qualitative components are based on established methodological approaches (e.g. group interviews in workshop format; [78]) to explore mechanisms, experiences and contextual influences, while quantitative components rely mainly on validated instruments and standardised questionnaires, which will be described later on in detail, to assess changes in the specified outcomes.

#### Sampling strategies and estimated sample sizes

This chapter describes the sampling strategies and estimated sample sizes for the implementation lead group, parents or caregivers as representatives for the primary school children and implementation facilitators within the evaluation design.

For the group interviews with the implementation lead groups, schools are selected using a purposeful maximum-heterogeneity sampling strategy to reflect the diversity of the overall *fit4future Kids* sample [79]. Sampling for the evaluation on the level of the implementation lead groups considers federal state, and a socio-economic index to ensure inclusion of schools from different educational contexts and with at least 25% from socioeconomically disadvantaged areas. For cohort 2 and cohort 3, this procedure yielded an initial target sample of 6 and respectively 10 primary schools (approximately one per federal state in Germany), which aligns with recommended ranges in in-depth qualitative research [80]. Schools for the group interviews were recruited via email and informed about the aims, procedure and the timetable of this evaluation aspect.

Furthermore, the evaluation targets the implementation lead group on a quantitative level, with a previous study indicating that approximately 80% of recruited schools will contribute data across the three cohorts at baseline and that attrition could remain at around 10% at subsequent timepoints [81]. Based on these response and attrition rates and the initially recruited 1,153 primary schools across the three cohorts, approximately *n =* 1,000 implementation lead groups complete the online-survey at the first measurement timepoint and 868 implementation lead groups complete every online survey in every of the following time points (estimated 472 complete cases in cohort 1 over five measurement time points). An a priori sample size analysis conducted with G*Power [82] indicated that this sample size for example in cohort 1 would be sufficient to detect small-to-medium effect sizes (f = 0.15, α = .05, Power = .80) in repeated-measures ANCOVA design testing for changes over time for example in the PDSA or the schools’ organisational health literacy by revealing a minimum cases of 424 for five measurement timepoints and a total of six covariate variables (school type, federal state, professional role, prior health promotion experience, existing guidelines for health promotion, readiness for implementing change).

Feedback surveys were administered to all implementation lead groups immediately following for example each workshop, with data aggregated across schools rather than analysed longitudinally per case. As the feedback survey aimed to ensure the quality of the programme no formal sample size planning was required because no longitudinal or inferential statistics were calculated. Usage data from the online platform was collected automatically for all participating implementation lead groups via cookies (opt-in), yielding descriptive analytics on engagement without predefined sampling targets. Implementation lead group participation rates were derived directly from attendance records for each workshop or peer-to-peer exchange session.

Assuming an average of approximately 250 children per German primary school [62] across all 1,153 participating schools, the total eligible parent or caregiver population is approximately 288,250 individuals. Based on response rates observed in comparable studies of around 10% [83,84] and accounting for an expected attrition rate of 90%, e.g. due to mismatching pseudonyms, school changes or graduation, for every measurement point [85], approximately 28,825 parents or caregivers could complete the survey at baseline and 2,882 parents or caregivers are expected to complete at least one follow up survey, with about 288 anticipated to participate at all three time points. For the longitudinal parent sample (n ≈ 288), an a priori sample size analysis in G*Power for a repeated-measures ANCOVA with three time points (f = 0.15, α = .05, Power = .80) and four control measures (age gender, SES and parental health status) indicate a minimum sample of 200 participants revealing that this sample size is sufficient to detect small-to-moderate within-subject effects, providing adequate power for the planned analyses [82].

The sampling of implementation facilitators follows a purposeful, information-oriented strategy [79] aimed at maximising variation in implementation experiences across the *fit4future Kids* programme. For each annual wave, we plan one focus group with approximately 6-10 facilitators, yielding an overall target sample of about 18–30 participants across three years. This range is consistent with established recommendations for in-depth focus group studies, which emphasise smaller groups and multiple sessions to enable rich interaction and comparative analysis [80]. The sample size is expected to be sufficient to identify recurring facilitator-level patterns, as well as contrasting cases, while allowing us to monitor when thematic saturation regarding typical implementation facilitators, barriers and response strategies is reached.

#### Detailed description of evaluation components

To provide an overview, Table 2 summarizes all evaluation components.

**Table 2.** Evaluation components.

| **Component** | **Data source / respondents** | **Frequency / timing** | **Focus / variables assessed** | **Level of analysis** |
| --- | --- | --- | --- | --- |
| Implementation lead group interviews | Implementation lead groups from selected schools | T0 – baseline,<br>T1 – mid-programme,<br>T2 – end of programme | Implementation experiences, barriers and facilitators, acceptability, feasibility, and sustainment | School |
| Implementation lead group online survey | Implementation lead groups | Baseline (T0) and annual follow-up surveys depending on cohort duration | HPS implementation, organisational health literacy, implementation determinants, organisational readiness for change, and organisational conditions | School |
| Programme feedback questionnaires | School personnel, parents or caregivers, and implementation facilitators | Immediately after workshops, peer-to-peer sessions, etc. | Acceptability, appropriateness, and feasibility of programme components | Individual |
| Usage data | Automatically collected usage metadata | Continuous throughout programme | Participation and engagement in programme components, including workshops, peer-to-peer exchange sessions, activity days, school coaching, and digital platform logins | School and programme |
| Parental/caregiver survey | Parents/caregivers as proxies for children | T0 – baseline, T1 – mid-programme, T2 – end of programme | Children's health behaviours and outcomes, family-level determinants, programme use, SES, parental health, and health literacy | Individual (child and parent/caregiver) |
| Implementation facilitators focus group interviews | Implementation facilitators | Once per year | Programme use, application of the PDSA cycle, school-level implementation processes, organisational changes, and facilitation needs | School and programme |

##### Group interviews with the implementation lead group

The group interviews with the implementation lead group aim to explore the programme’s implementation, perceived barriers and facilitators, acceptability and feasibility, adapting processes, and sustainment. They are conducted with implementation lead groups’ from a subset of schools that agreed to participate (n = 4 in cohort 2 and n = 6 in cohort 3). Data collection takes place at three time points: baseline, after one year, and at the end of the programme after one or two years depending on the cohort. The semi-structured interview guideline was developed by the scientific partners and builds on the CFIR [43,44], relevant literature [86], and established approaches for qualitative research [80]. Further, the interview was structured in an interactive workshop-like way to encourage active reflection [78]. An overview of the procedure is provided in S10.

##### Implementation lead group online survey

The online survey for the implementation lead group was developed by the scientific partners consisting of psychometrically approved and widely used questionnaires. In the pretesting phase, the survey was discussed in six peer group meetings with school personnel using cognitive interviewing techniques [87]. In the evaluation process, the online survey is completed either collectively or by one representative of the implementation lead group annually depending on the cohort and respectively the duration of the programme.

The first part of the survey concerns descriptive questions, such as the role of the survey participant within the school (e.g. teacher, principal, other personnel), prior participation in other health promotion programmes, or the school’s curriculum concerning school-based all-day programmes and extracurricular activities.

Then the participants reported their school’s HPS framework implementation by completing the HPS questionnaire, which is grounded in the German evaluation guidelines for school-related health promotion issued by the statutory health insurance funds [52]. The instrument is conceptually linked to the PDSA cycle and consists of 19 items covering several key dimensions of school health promotion, including the planning, implementation and evaluation of health promotion activities, the current status of ongoing measures, the development and use of supportive structures, the integration of health topics into teaching, and the creation of a supportive school environment. Items are rated on four- or five-point rating-type scales (e.g. “not planned yet”, “in planning”, “implementation started”, “partially achieved”, “fully achieved”). An example item targeting structural aspects is: “Our school has established structures that support the planning and implementation of health promoting activities.” An overall HPS implementation score is computed by summing all item responses ranging from 0 to 76 with higher values reflecting a higher degree of HPS implementation. In a preliminary data analysis, the scale has demonstrated good structural validity within a confirmatory factor analysis and excellent internal consistency, with Cronbach’s alpha reported at α = 0.91.

Schools’ organisational health literacy is assessed using the Organisational Health Literacy of Schools Questionnaire in a Short Form (OHLS-Q-SF; [88]). The OHLS-Q-SF captures the eight standards (include health literacy into the school’s mission statement, health literacy as part of school development, promote and enhance health literacy in daily school life, health literacy of students, health literate school staff, health literate communication at school, enhance health literacy in the school environment, networking and cooperation) of a health-literate school with a total of 8 items, each rated on a four-point Likert scale (“strongly disagree”, “disagree”, “agree”, “strongly agree”). A sum score for schools’ health literacy will be calculated by adding all item responses (range 0–24), with higher scores indicating a higher level of organisational health literacy at school [89]. The questionnaire showed good validity and reliability, which was tested amongst others by examining construct validity and with an acceptable Cronbach’s alpha (α = 0,79; [90]).

Furthermore, to examine the school’s likelihood of implementing the programme, the organisational readiness for implementing school-level changes will be assessed using an adapted German version of the Organisational Readiness for Implementing Change (ORIC) measure [91,92]. The original ORIC is a 10-item, theory-based self-report instrument designed to capture staff members’ collective commitment to, and perceived capability for, implementing a specific organisational change and has shown a one-factor structure with excellent internal consistency in German (Cronbach’s α = 0.93). For the present study, the wording of the items was transferred from the hospital to the school context (for example, by replacing references to the clinic with references to the school and specifying the targeted change), while retaining the 5-point Likert response format ranging from “disagree” to “agree”. Following the authors’ recommendation, we use the unidimensional German version and compute a total readiness score by summing all items, with higher scores indicating a higher level of organisational readiness to implement the intended school-related changes [91]. The adapted version of the ORIC demonstrated good construct validity and excellent internal consistency in preliminary analysis (Cronbach’s α = 0.93).

Lastly, members of the implementation lead group completed a brief scale capturing perceived organisational conditions for implementing and sustaining school health promotion. The eight items were developed inductively based on qualitative workshops with implementation lead group members and target central aspects discussed there, including financial and infrastructural support (e.g. availability of funding and suitable spaces), personnel resources, internal communication structures, the lived school mission regarding health promotion, and active support from teachers and parents, as well as the continuous use of a structured health promotion process [93]. Items are rated on a 5-point Likert scale and can be summarised into an overall score, with higher values indicating more favourable organisational conditions for school health promotion. In the present sample, the scale showed acceptable internal consistency (Cronbach’s alpha = 0.78). A preliminary one-factor solution suggested that the items share common variance but also reflect partly distinct facets of organisational support; thus, analyses will primarily use the total score while reporting item-level results exploratorily.

##### Programme feedback questionnaires

The feedback questionnaires aim to capture immediate participant responses on the acceptability, appropriateness, and feasibility of programme components. They are administered to school personnel and parents or caregivers after workshops (peer-to-peer exchange sessions or workshops) as well as to implementation facilitators after their training sessions. Data are collected via an online survey following each event. Items are self-developed and based on the CFIR antecedent outcomes assessment [45]. Questionnaires primarily use scaled items and also include open-ended questions to allow for additional feedback.

##### Usage data

Usage data aim to monitor objective implementation outcomes. They are derived from meta-data including participation in peer-to-peer exchange sessions, workshops, activity days, as well as requests for school coaching and number of logins to the digital platform. The data collection on these objective implementation outcomes is done continuously throughout the health promotion programme with data being exported for example on the number of logins to the digital platform every six months for all participating schools. These objective implementation outcomes complement the self-reported quantitative and qualitative data to enable triangulation of implementation outcomes [94].

##### Parental or caregiver survey

The parental or caregiver survey is an online questionnaire to assess children’s health behaviours and outcomes, family-level determinants such as, parental health literacy, family health climate, and the use of *fit4future Kids* programme components. It is administered in cohort 2 at baseline, after one year, and after two years. The survey was developed in line with the German statutory health insurance funds’ guidelines for the evaluation of health promotion [52] covering amongst sociodemographic children-related characteristics, socioeconomic status of parents and caregivers, parental health and health literacy the central health areas of physical activity, dietary behaviour, mental health and media use on a family health climate, a behavioural and an affective perspective. All scales used are implemented in large cohort studies or derive from reliable and valid questionnaires, which have found widespread acceptance in the research on health promotion in the family and school context.

The parental survey includes basic sociodemographic characteristics (e.g., child’s age, sex assigned at birth, household size, languages spoken at home) and anthropometric data (height and weight), enabling the calculation of BMI z-scores using WHO reference standards [95]. Implementation-related parameters are captured via self-developed items on familiarity with *fit4future Kids* and participation in key programme components (e.g., parent evenings, online seminars, use of the health platform). Together, these measures allow for a comprehensive evaluation of programme effects on child and family outcomes and the role of family-level determinants and context conditions.

Parental SES is measured in line with the KiGGS SES module [96]. Parents report highest general school and vocational qualifications (for each parent), current employment status, occupational position (including supervisory responsibility), and total monthly net household income. Based on these indicators, a standardized SES index combining education, occupation, and income is constructed, with higher scores indicating higher SES. The index has demonstrated strong validity in prior studies through robust gradients in morbidity and health-related behaviours [96].

Parental health status is assessed with a single global item: “How would you describe your health in general?”. Response options are “very good”, “good”, “moderate”, and “poor”, and can be coded on a four-point scale such that higher values reflect better parental health. This type of single-item measure has repeatedly been shown to be a valid indicator of overall health status in large population-based studies [96,97].

Parental health literacy is assessed using four items from the “health promotion” domain of the short form of the European Health Literacy Survey Questionnaire (HLS-EU-Q16) [98]. The items tap parents’ perceived ease of finding, understanding, appraising, and applying health-related information. A sample item is: “In your opinion, how easy is it to find information about behaviours that promote your mental well-being?”. Responses are given on a 4-point Likert scale from “very difficult” to “very easy”. Item scores are summed to create an index ranging from 0 to 16, with higher scores reflecting higher parental health literacy. A previous study has reported satisfactory internal consistency (Cronbach’s α ≥ 0.80) and supporting associations of this HLS-EU-Q16 domain with education, health behaviours, and health status, supporting reliability and validity [98].

The family health climate regarding physical activity and nutrition is measured using short forms of the Family Health Climate Scale (FHC) [99]. The physical activity subscale (FHC-PA) includes 14 items capturing the dimensions of value, cohesion and information of a physically activating family climate (e.g., “…we agree that physical activities are part of daily life.”). The nutrition subscale (FHC-NU) comprises 17 items on the dimensions of value, cohesion, communication, and consensus addressing the importance of healthy eating, agreement on nutrition, and shared meals (e.g., “a healthy diet plays an important role in our lives.”). Items are rated on 4-point rating scales from 0 = ‘definitely false’, 1 = ‘rather false’, 2 = ‘rather true’, 3 = ‘definitely true’. Subscale scores are computed by summing item responses; higher scores indicate a more supportive family health climate in the respective domain. The original validation study reported good to excellent internal consistency and meaningful correlations with dietary and activity patterns, supporting reliability and construct validity [99].

Family health climate on a social-emotional level is examined with the corresponding subscale of the Family Health Scale (FHS) concerning social-emotional health processes [100]. The 13 items capture emotional connection, mutual support, shared problem-solving, hope, and meaning within the family (e.g., “There is a feeling of togetherness.”). Parents respond on a 5-point Likert scale ranging from “strongly disagree” to “strongly agree”. Item scores are summed to form a total social-emotional family health score, with higher scores indicating healthier social-emotional family processes. The FHS has demonstrated very high internal consistency (Cronbach’s α = 0.92), a clear factor structure, and measurement invariance indicating good reliability and construct validity [100].

Family-level media competence is assessed using six items adapted from the DIVSI U9 study [101]. These items assess parents’ perceived knowledge and confidence regarding their child’s media use (e.g., “I feel well able to teach my child how to use digital media and the internet safely.”) on 4-point Likert scales. Summed scores yield an index of familial media competence, with higher scores reflecting greater perceived competence. Due to the lack of alternative validated questionnaires, we employed this currently unvalidated questionnaire, which was used in the large German cohort study on media use in children without reporting psychometric properties [101].

Children’s physical activity is assessed using two items adapted from the KiGGS and MoMo questionnaires [102], asking on how many days in a usual week and in the previous week the child was physically active for at least 60 minutes per day. Response options range from 0 to 7 days. These items are used separately and combined as indicators of habitual and recent moderate-to-vigorous physical activity, with higher values reflecting higher activity levels. Previous work has shown acceptable validity e.g. through associations with objective activity measures and fitness [102].

Children’s dietary behaviour is captured by three frequency items adapted from KiGGS [15] assessing the consumption of fruit, cooked vegetables, and raw vegetables/salad in the past weeks (e.g., “How often has your child eaten a portion of raw vegetables or salad?”). Response categories range on a 10-point rating scale from “never” to “more than five times a day”. These items are used to derive frequency indicators and combined fruit-vegetable scores, where higher values reflect a more favourable dietary pattern. The items have been widely used in population-based surveys and show plausible gradients with health and weight outcomes [15].

Screen-based media use is assessed following KiGGS and ScreenQ conventions with a total of six items [102,103]. Parents report the average daily time their child spends with television/videos, game consoles, and internet-enabled devices (computer, smartphone, tablet) on a 6-point rating scale ranging from “not at all” to “more than 4 hours per day”. Three additional items derived from the ScreenQ assess whether media are used alone or together with an adult and how frequently parents co-view or discuss content. From these items, composite indicators of total screen time and parental co-regulation are derived, with higher screen-time scores reflecting greater daily exposure. Prior studies have documented acceptable reliability and criterion validity of ScreenQ-based measures e.g. in relation to outcomes of emergent literacy and expressive vocabulary [103].

Affective responses to physical activity were assessed using a parent-adapted short version of the Physical Activity Enjoyment Scale (PACES-S) [104]. The scale consists of four items that capture the extent to which the child is perceived as enjoying physical activity (e.g., “Being physically active is enjoyable for my child.”). Parents rate their agreement on a five-point Likert scale ranging from “strongly disagree” to “strongly agree.” A total score is computed, with higher values indicating greater enjoyment of physical activity. The short PACES has demonstrated solid psychometric properties, including good internal consistency in a previous study (α = 0.82) and evidence for concurrent validity through positive associations with both self-reported and device-measured physical activity levels [104].

Eating-related affect and preferences are assessed with a short form of the Enjoyment of Food subscale from the Child Eating Behaviour Questionnaire (CEBQ) [105,106]. Seven items capture positive attitudes towards eating and interest in food (e.g., “My child loves food”, “My child likes to try new foods.”), rated on a 5-point Likert scale from “never” to “always”. Summed scores yield an index of positive eating-related affect and openness to foods, with higher scores reflecting greater enjoyment of and interest in food. Previous applications of the CEBQ have demonstrated acceptable to good internal consistency (α ≥ 0.73) and stable factor structures across diverse samples [105].

Problematic media use is measured using the short form of the Problematic Media Use Measure (PMUM-SF) [107]. Nine items capture addictive-like patterns of screen use (e.g., “It is hard for my child to stop using screen media.”, “My child gets frustrated when not allowed to use screen media.”), rated on a 5-point scale from “never” to “always”. Item scores are summed to form a total problematic media use score; higher values indicate more pronounced problematic use. The PMUM-SF has shown excellent internal consistency (α = 0.90), a unidimensional factor structure, and convergent validity through associations with behavioural problems and media-related family conflict [107].

Children’s health-related quality of life is assessed via the parent-proxy KINDL [108]. The 24 items cover six subscales (physical well-being, emotional well-being, self-esteem, family, friends, school), rated on a 5-point scale (“never” to “always”) for the past week (e.g., “In the past week, my child felt comfortable at home.”). Scores are transformed to 0-100 with higher scores indicating better well-being. The KINDL demonstrates good internal consistency (α = 0.80) and robust construct/criterion validity across samples and finds wide usage to examine health-related quality of life in children [108,109].

##### Implementation facilitators focus group interviews

Implementation facilitator focus group interviews are conducted once per year (summer 2024, 2025, 2026), either face-to-face or via video call. Each interview session includes 6-10 implementation facilitators who are responsible for schools from at least two different German federal states to avoid region-specific bias and to stimulate exchange across educational contexts. A semi-structured interview guide was constructed in advance and piloted with two facilitators. Its development followed established procedures for qualitative interviewing and focus group research, including Helfferich’s stepwise interview planning [80]. The guide contains a fixed set of main questions and standardised prompts; all groups work through the same questions in the same order. In the interviews, implementation facilitators are explicitly asked to speak about the schools they supervise rather than about themselves. Four domains are covered: (1) use and non-use of specific *fit4future Kids* components (e.g. workshops, peer sessions, materials) and reasons for this; (2) concrete examples of how schools apply the PDSA cycle; (3) observable changes in schools’ health-promoting activities and organisational routines over the past school year; and (4) situations in which additional facilitation was needed and how it was provided. For each domain, facilitators are asked to name both schools that progressed well and schools that struggled, and to give illustrative examples.

#### Data management

The evaluation is conducted in compliance with the General Data Protection Regulation (GDPR) and ethical guidelines, approved by the Ethics Committee of the Technical University of Munich (TUM) prior to the first data collection (2023-564-S-NP, 2023-12-NM-KH). In addition, the study was registered retrospectively on the Open Science Framework (https://doi.org/10.17605/OSF.IO/CM6FK). Collected data is used exclusively for stated programme evaluation, refinement or research purposes and treated confidentially. Data within the schools is collected in a pseudonymised form and all individuals participating in the survey complete consent forms with separate saving locations for data and consent forms of participating schools. Data is stored on the servers of the Leibniz Computing Center (Leibniz Rechenzentrum, LRZ) of the Bavarian Academy of Sciences for a maximum of 10 years after the evaluation ends. No data is shared with any other institution. Participation in the evaluation is voluntary, and participants can withdraw their consent at any time, immediately ending data collection from that moment on. Participants have the right to access their data and request correction, restriction, deletion or transfer of their data.

#### Data analysis

All qualitative data (group interview workshops with school implementation lead groups and focus group interviews with implementation facilitators) will be analysed using a rigorous, multi-step approach that combines the framework method with thematic analysis [110]. All sessions are audio-recorded and transcribed verbatim. Field notes taken during and immediately after each session document group dynamics and contextual circumstances (e.g. technical problems, time pressure, dominant speakers) are used to enrich interpretation of the transcripts.

In a first step, transcripts will be imported into MAXQDA and coded using an a priori codebook based on the updated CFIR, which was previously adapted to the school context in this study and refined through iterative test coding and double-coding procedures. Two researchers will independently apply the analytical framework to all new transcripts, discuss discrepancies, and agree on a final coding scheme. For each CFIR construct, data will then be charted into a framework matrix by school or stakeholder group, and valence ratings (facilitator, barrier, mixed/ambivalent) will be assigned. As in Sterr et al., these ratings will be synthesized and visualized in a colour-coded matrix heat map to support transparent cross-case comparison and to identify contextual tendencies or configurations that may be relevant for implementation [93].

In parallel, and particularly for the implementation facilitator focus groups, we will conduct thematic analysis as described by Clarke and Braun [110]. This involves (1) familiarization with the data, (2) generating initial inductive and deductive codes, (3) collating codes into candidate themes, (4) reviewing and refining themes against the coded extracts and the full data set, (5) defining and naming themes, and (6) producing analytic narrative accounts that integrate thematic patterns with the quantitative and process data. Coding will be conducted by at least two researchers, with regular peer debriefings to reflect on assumptions, resolve disagreements, and enhance reflexivity [110].

Across data sources, qualitative findings will be used to (a) assess schools’ progress and challenges in applying the PDSA cycle and other programme components from an external vantage point, (b) identify recurrent implementation facilitators, barriers, and response strategies that are not readily captured by questionnaires, and (c) generate hypotheses about contextual configurations that may be associated with differential implementation trajectories. Where appropriate, qualitative themes and CFIR-based matrix patterns will be integrated with quantitative indicators (e.g. school-level readiness scores, implementation fidelity, and outcome trajectories) to inform mixed-methods interpretation and to guide the refinement of implementation support strategies.

Quantitative analyses will be conducted using RStudio (2026.04.0, Build 526). Data preparation will involve plausibility checks, harmonisation across cohorts and measurement waves, range validations, and variable recoding. Derived measures such as WHO BMI-z-scores will be calculated according to established growth reference standards [95,111].

Missing data will be examined descriptively and addressed primarily using k-nearest-neighbour imputation, a pragmatic, non-parametric method that performs well when data are plausibly Missing at Random [112,113]. To assess robustness, sensitivity analyses will include (a) multiple imputation using chained equations [114], (b) complete-case analyses, or (c) full-information maximum likelihood estimation within multilevel SEM, consistent with recommendations for handling missingness in clustered longitudinal data [115].

All multi-item scales will be computed following published scoring guidelines. Internal consistency will be assessed using Cronbach’s α and McDonald’s ω, acknowledging the limitations of α and the advantages of ω for multidimensional constructs [116,117]. Where sample size permits, longitudinal measurement invariance across waves and cohorts (configural, metric, and scalar) will be tested via confirmatory factor analysis to ensure that changes over time reflect true score changes rather than measurement artefacts [118].

Descriptive analyses will summarise child, parent, and school characteristics. Response rates and attrition patterns will be compared descriptively across cohorts, following guidelines for reporting observational clustered data [119].

Given the hierarchical structure of the data, primary analyses will employ linear mixed-effects models with random intercepts for schools, following best practices for clustered and longitudinal health promotion data [120,121]. Fixed effects will include measurement wave, cohort, and federal state; continuous predictors will be standardised to facilitate interpretation. When justified by intraclass correlations and variance partitioning, random slopes (e.g. for time) will be added [122]. Baseline-adjusted longitudinal models (ANCOVA logic) will be used for change estimation, a strategy known to increase statistical power and reduce bias in non-randomised longitudinal studies [123].

Primary outcomes include amongst others parent-proxy reported children’s health-related quality of life and health behaviour on a personal level as well as school health literacy on an organisational level. Secondary outcomes include parental health literacy, family health climate, and school-level indicators such as HPS implementation and organisational readiness for implementing change. Standardised coefficients with 95% confidence intervals will be reported, following recommendations for effect estimation in mixed-effects modelling [124]. Moderation analyses will examine differential effects based on socioeconomic status, and baseline organisational readiness, reflecting PROGRESS-Plus equity considerations [125].

Objective implementation exposure will be analysed using usage metrics (e.g., platform logins, workshop participation, peer-to-peer session attendance). These variables will be incorporated either as time-varying covariates or as composite engagement indices to link implementation dose with changes in outcomes, consistent with RE-AIM and implementation outcomes frameworks [75,77]. Analyses will adjust for baseline covariates to reduce self-selection bias.

Exploratory analyses will use coincidence analysis (CNA) to identify minimally sufficient and necessary configurations of contextual conditions and implementation processes. CNA will be conducted following established methodological standards [126]. Calibration thresholds, consistency and coverage metrics, and robustness checks will be reported transparently.

Robustness checks will include alternative missing-data approaches, complete-case re-estimation, different model specifications (e.g., random intercept vs. random slope), and alternative link functions for skewed or discrete outcomes, following recommendations for model diagnostics in applied multilevel research [127].

A theory-driven two-level structural equation model will be estimated using robust maximum likelihood (MLR) with clustering by school, enabling simultaneous testing of hypothesised pathways at the individual and school level [128]. Model fit will be evaluated using robust CFI, TLI, RMSEA, and SRMR [129,130]. Explained variance (R²) will be reported separately for Level 1 and Level 2 outcomes.

All analyses will adhere to STROBE-Cluster and StaRI reporting guidelines [131,132]. Code, variable derivations, and outputs will be version-controlled and archived on the Open Science Framework. Deviations from the preregistered analysis plan will be documented.

## Discussion

In this protocol, the *fit4future Kids* programme is presented as a nationwide multi-component school-based intervention in Germany that aims to promote children’s physical activity, nutrition, mental health, and digital media use, while simultaneously strengthening primary schools’ capacity for sustainable health promotion. The intervention aligns with the HPS framework by the WHO [26] and integrates evidence-based components that have previously been identified as effective in school-based health promotion programmes [23,32,133,134].

Beyond the content of the intervention itself, the implementation of *fit4future Kids* is informed by established implementation science frameworks, particularly the CFIR [43,44].In addition, behaviour change models such as the COM-B model [50] and BCTs [51] are incorporated to support the translation of health promotion strategies into practice. By combining these frameworks, the programme seeks to address well-documented implementation challenges commonly observed in school-based interventions, including variability in school contexts, limited resources, and competing priorities [39].

Although *fit4future Kids* may appear comprehensive and potentially complex, the programme is deliberately designed to allow flexibility for participating schools. Schools can adapt and tailor health-promoting activities to their specific needs and engage with different forms of implementation support. Such adaptable implementation strategies have been identified as particularly relevant and frequently requested within school settings [47,93].

The evaluation concept is equally comprehensive and follows a mixed-methods approach involving multiple target groups. This design allows the integration of quantitative and qualitative perspectives and facilitates the triangulation of findings across different data sources [94]. Moreover, the large and diverse sample covering approximately 9% of German primary schools, with around 25% located in socioeconomically disadvantaged areas provides a valuable opportunity to explore heterogeneous implementation trajectories and to assess potential equitable effects on child health behaviours and outcomes.

Despite these considerable strengths, several limitations should be acknowledged. First, the study design does not include a control group, which limits the ability to draw causal conclusions regarding programme effects. Nevertheless, alternative analytical approaches, such as CNA, may help to identify causal pathways and combinations of conditions associated with successful outcomes. Second, the large-scale nature of the programme may limit the extent to which individualized support, supervision, and detailed monitoring of implementation can be provided across all participating schools. Third, the flexibility granted to schools regarding which components they implement may make it difficult to attribute observed changes in child health behaviours to specific intervention elements. Finally, the evaluation primarily relies on proxy reports (e.g. from parents, teachers, and school staff) and does not directly capture children’s own perspectives. This may limit insights into children’s subjective experiences, perceptions, and acceptance of the programme, which are important for a comprehensive understanding of its effectiveness.

Furthermore, outcome assessment at the child level relies partly on parental proxy reports, which may introduce reporting bias. Data collection across both school and family contexts primarily relies on self-reported measures, which may also increase the risk of response bias. Finally, long-term follow-up beyond the intervention period is only available for the first and second programme cohorts, limiting the ability to examine sustained behavioural changes and structural implementation effects for the third cohort.

Taken together, the *fit4future Kids* programme provides a large-scale real-world example of how evidence-based health promotion and implementation science frameworks can be combined to support sustainable health promotion in primary schools. The evaluation results are expected to generate important insights into the implementation and effectiveness of multi-component school-based health promotion programmes and may inform future large-scale initiatives aimed at improving child health and reducing health inequalities.

## Conclusion

Overall, *fit4future Kids* exemplifies an evidence-informed, multi-component school-based intervention promoting health behaviours while building organisational capacity for sustainable health promotion. By integrating theory-driven frameworks, inclusive implementation strategies and a multi-level evaluation design, it provides a replicable model for health promotion primary schools. Transparent documentation of intervention and evaluation processes contributes to advancing best practices in international school health promotion and informs future efforts to address childhood NCD risk holistically and sustainably.

## Data Availability

No datasets were generated or analysed during the current study. All relevant data from this study will be made available upon study completion.

## Supporting information

S0: TIDieR checklist

S1: Overview of programme components with Content, COM-B, BCTs and SISTER mapping

S2: Training concept for implementation facilitators

S3: Idea Box (digital repository)

S4: Blog (How-To, Best-Practice, inspirational posts)

S5: Useful information (program-level materials and PDSA tools by cohort)

S6: *fit4future box* (play and sports equipment)

S7: *fit4future box* (action cards)

S8: Workshops for implementation lead groups

S9: On-site Health Activity Days

S10: Group interview-guideline (evaluation)

